# Use of alternative RNA storage and extraction reagents and development of a hybrid PCR-based method for SARS-CoV-2 detection

**DOI:** 10.1101/2020.11.21.20236216

**Authors:** Julie Yang, Elias Salfati, Damian Kao, Yuliana Mihaylova

## Abstract

The COVID-19 pandemic has presented multiple healthcare challenges, one of which is adequately meeting the need for large-scale diagnostic testing. The most commonly used assays for detection of SARS-CoV-2, including those recommended by the Center for Disease Control and Prevention (CDC), rely on a consistent set of core reagents. This has put a serious strain on the reagent supply chain, resulting in insufficient testing. It has also led to restricted animal testing, even though there are now multiple reports of animals, particularly cats, ferrets and minks, contracting the disease. We aimed to address the diagnostic bottleneck by developing a PCR-based SARS-CoV-2 detection assay for cats (and, potentially, other animals) which avoids the use of most common reagents, such as collection kits optimized for RNA stabilization, RNA isolation kits and TaqMan-based RT-PCR reagents. We demonstrated that an inexpensive solid-phase reversible immobilization (SPRI) method can be used for RNA extraction from feline samples collected with DNAGenotek’s ORAcollect RNA OR-100 and PERFORMAgene DNA PG-100 sample collection kits, optimized for RNA or DNA stabilization, respectively. We developed a dual method SARS-CoV-2 detection assay relying on SYBR RT-PCR and Sanger sequencing, using the same set of custom synthesized oligo primers. We validated our test’s specificity with a commercially available SARS-CoV-2 plasmid positive control, as well as two in-house positive control RNA samples. Our assay’s sensitivity was determined to be 10 viral copies per reaction. Our results suggest that a simple SPRI-dependent RNA extraction protocol and certain sample collection kits not specifically optimized for RNA stabilization could potentially be used in cases where reagent shortages are hindering adequate COVID-19 testing. These ‘alternative’ reagents could be used in combination with our COVID-19 testing method, which relies on inexpensive and readily available SYBR RT-PCR and non-fluorescent PCR reagents. Depending on the detection goals and the laboratory setup available, the SYBR RT-PCR method and the Sanger sequencing based method can be used alone or in conjunction, for improved accuracy. Although the test is intended for animal use, it is, in theory, possible to use it with human samples, especially those with higher viral loads.

## Introduction

The COVID-19 pandemic has put an unprecedented strain on almost every country’s healthcare system. In addition to insufficient hospital beds and personal protective equipment for healthcare workers, this pandemic has also been marked by a shortage of COVID-19 diagnostic testing. The most frequently used assay type, the SARS-CoV-2 RT-PCR, relies on the same set of preferred core reagents. For this type of test, the CDC recommends the use of commercially available RNA isolation kits and RT-PCR reagents designed for use with an oligo probe (i.e., the TaqMan approach)^1^. This has led to multiple reports of RNA isolation kit shortages^2^, slowing down testing efforts at a time when testing speed is crucial. In addition, from our own internal observations, many TaqMan-based RT-PCR reagents, as well as nasopharyngeal and oropharyngeal swab kits optimized for RNA stabilization, were backordered and/or experienced a price increase at some point during the pandemic. These supply chain bottlenecks have also led to very sparse testing of animals suspected to have COVID-19, despite the fact that there has been strong research evidence indicating that animals (particularly cats, ferrets and minks) can contract and spread COVID-19 to other animals^3,4,5^. Even with limited testing, there are currently over 60 reported COVID-19 positive animal cases in the US alone, as catalogued by the United States Department of Agriculture^6^. Recently, COVID-19 diagnostic tests designed specifically for animal use were made available for clinical use only^7^. However, these tests still rely on the same set of core reagents and, therefore, do little to address or circumvent reagent shortages. To address this issue, we developed a SARS-CoV-2 PCR-based test for use in cats (and potentially other animals) that does not rely on the reagents most typically used in SARS-CoV-2 RT-PCR tests. Our assay can be used with a sample collection kit optimized for DNA stabilization or RNA stabilization and does not use commercial RNA extraction kits or TaqMan based RT-PCR reagents. Our test is comprised of two different workflows - SYBR RT-PCR and Sanger sequencing. These can be used together or separately, depending on the available reagents and lab setup. The availability of such a test would be important for improving our understanding of the spread and impact of the SARS-CoV-2 virus in animals.

## Results

### The PERFORMAgene PG-100 DNA collection kit from DNAGenotek can be used for RNA stabilization and isolation

We collected an oropharyngeal swab sample from the same cat using two different swab collection kits - the ORAcollect RNA OR-100 and the PERFORMAgene DNA PG-100 (both available from DNAGenotek), optimized for RNA and DNA stabilization, respectively. We then extracted total RNA from each of these samples using a published method for purifying nucleic acids by solid-phase reversible immobilization (SPRI)^8^, which does not rely on a commercial kit. Gel electrophoresis indicated that we were successful in extracting total RNA from each of the two samples (**Figure 1A**). Quantification of the extracted RNA suggested that our extraction protocol yielded higher quantities of RNA from the sample collected with the PERFORMAgene DNA PG-100, compared to the sample collected with the ORAcollect RNA OR-100 (**Figure 1B**). RNA purity was quantified by absorbance measurements taken at 260/280 nm and 260/230 nm. Our data suggest that the RNA extracted from the ORAcollect RNA OR-100, although lower quantity, is of better quality. However, the best indicator of RNA quality is, ultimately, its functionality in a particular application of interest. We wanted to investigate whether the RNA extracted from the PERFORMAgene DNA PG-100 sample contained feline mRNA and viral RNA. We successfully detected two housekeeping genes (GAPDH and RSP19) in each of the two samples, although RSP19 was of a lower quantity in the RNA extracted from the PERFORMAgene DNA PG-100 sample compared to the RNA extracted from the ORAcollect RNA OR-100 sample (**Figure 1C**). Therefore, while both collection kits allowed successful extraction of feline mRNA from an oropharyngeal sample, our results indicate that some feline mRNAs might have different abundance between the two collection kits.

**Figure 1.**
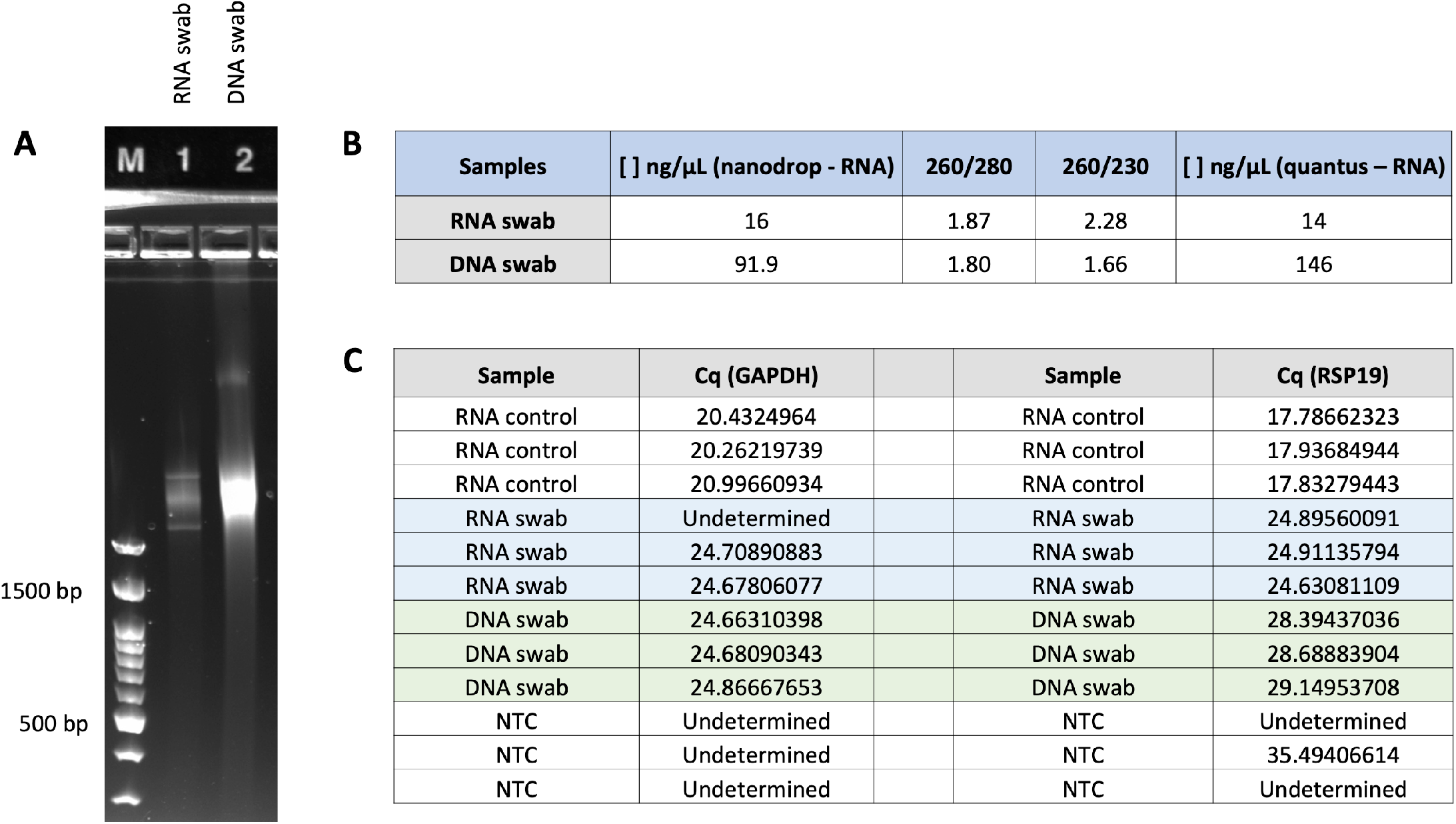
Yield and quality of RNA extracted from a feline oropharyngeal swab sample collected with the ORAcollect RNA OR-100 (RNA swab) and the PERFORMAgene DNA PG-100 (DNA swab). (A) Gel electrophoresis comparing the total RNA extracted from each sample collection kit. The M lane contains a nucleic acid size marker. (B) Measures of RNA amount and purity using ThermoFisher Scientific NanoDrop and Promega Quantus spectrophotometers. (C) RT-PCR results quantifying the expression of two housekeeping genes (GAPDH and RSP19) in the RNA extracted from each kit.

We next tested whether the total RNA extracted from the PERFORMAgene DNA PG-100 sample contained viral sequences. We prepared an RNA-seq library from the extracted RNA and ran the sequencing results through a microbial sequence classifier to identify 33 viral genomes, at least 9 of which had RNA genomes (**Table 1**). Surprisingly, despite the DNAse treatment step in the RNA-seq library preparation process, we still detected some viruses with DNA genomes in the sample.

**Table 1.** High, medium and low abundance viral sequences detected in the RNA extracted from the PERFORMAgene DNA PG-100 sample. Viruses with known RNA based genomes are marked in orange.

| High abundance viruses | Medium abundance viruses | Low abundance viruses |
| --- | --- | --- |
| Coliphage phi-X174 | Tomato brown rugose | Haemophilus phage HP1 |
| Puma feline foamy | Enterobacteria phage lambda | Haemophilus phage HP2 |
| Feline foamy virus | Enterobacteria phage HK630 | Campylobacter phage CP21 |
|  | Oryza sativa endornavirus | Woolly monkey sarcoma virus |
|  | Staphylococcus phage SPbeta-like | Enterobacteria phage M13 |
|  | RD114 retrovirus | Enterobacteria phage T7 |
|  | Enterobacteria phage DE3 | Tokyovirus A1 DNA |
|  | Brome mosaic virus | Pseudomonas phage pf16 |
|  | Human endogenous retrovirus | Aeromonas phage pf16 |
|  | Proteus phage VB_PmiS-Isfahan | Aeromonas phage 4_4572 |
|  | Pandoravirus dulcis | Picornavirales Bu-3 gene |
|  | Gallid herpesvirus 2 | Arthrobacter phage Glenn |
|  | Phytophthora infestans RNA | Arthrobacter phage Korra |
|  | Propionibacterium phage Pirate |  |
|  | Paramecium bursaria Chlorella |  |
|  | Pandoravirus salinus |  |

Our results suggest that the PERFORMAgene DNA PG-100 sample collection kit has the potential to be used for preservation of both host mRNA and viral RNA. This makes it a tool that merits further investigation in a SARS-CoV-2 diagnostic testing setting, especially in situations where collection kits optimized for RNA stabilization are in short supply.

### A dual method SYBR RT-PCR and Sanger sequencing approach can be used for detection of SARS-CoV-2

In an attempt to avoid using TaqMan RT-PCR reagents for SARS-CoV-2 detection, we tested whether a combination of SYBR RT-PCR and Sanger sequencing could deliver reliable detection of the virus. We designed oligo primers amplifying a 328 bp sequence in the N gene region of the SARS-CoV-2 genome and looked at the conservation between the homologous target regions of SARS-CoV-2, SARS-CoV and the Feline enteric coronavirus (FECV) (**Figure 2**). The FECV sequence is sufficiently divergent as to not present a threat for viral cross-detection. SARS-CoV, however, shares high homology with SARS-CoV-2 and our primer pair would work equally well for amplifying either of the two viral sequences. Therefore, the SYBR RT-PCR part of our assay, when used in isolation, cannot distinguish between the two viruses. The Sanger sequencing component of our test, on the other hand, can clearly distinguish between SARS-CoV and SARS-CoV-2 due to the single nucleotide resolution that it provides (**Figure 3**).

**Figure 2.**
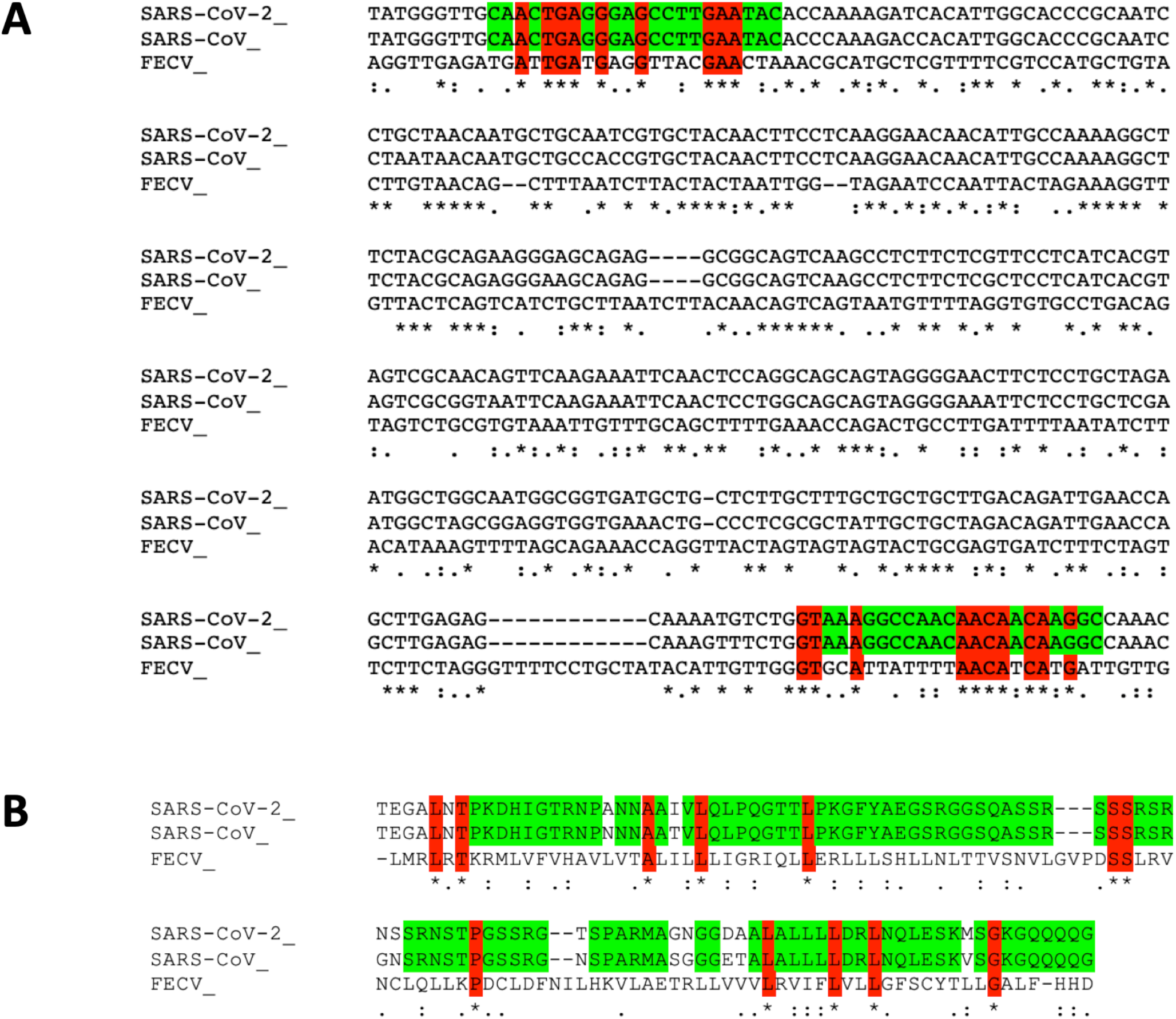
Sequence alignment between the SARS-CoV-2, SARS-CoV and FECV viral genomes, focusing on the corresponding region targeted for amplification in our assay. (A) Nucleotide alignment between the 3 viruses, with the primers used in our assay shown in color; red indicates nucleotides that are the same between all three viruses and green indicates nucleotides shared only between SARS-CoV-2 and SARS-CoV. (B) Protein alignment of the whole 328 bp region targeted for amplification in our assay; color code is the same as in (A).

**Figure 3.**
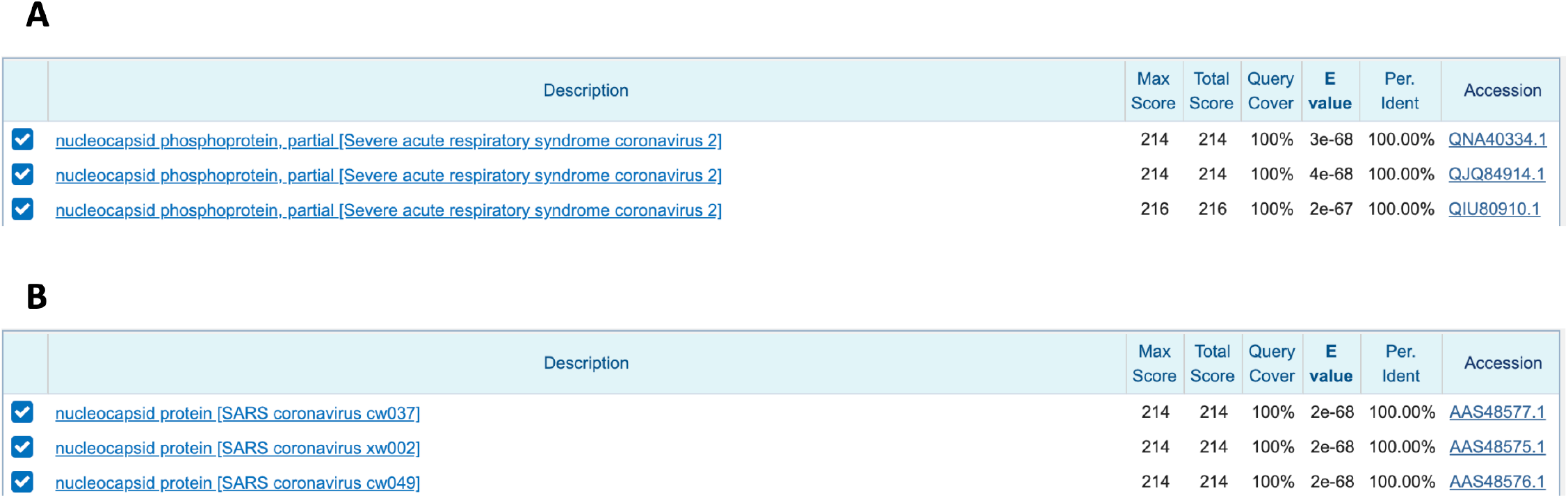
Top 3 results when (A) the SARS-CoV-2 or (B) the SARS-CoV region targeted for amplification by our primers is used as input into NCBI’s BLASTp tool.

We tested the above primers in our SYBR RT-PCR and Sanger sequencing protocols (**Figure 4**). We used a plasmid containing the N gene region as a spike-in synthetic positive control in the feline cDNA used in our PCR reactions. Gel electrophoresis showed a clear and specific amplification signal of the expected size (328 bp) in the positive control samples when the reaction was spiked-in with 1000 or 500 viral copies, respectively (**Figure 4A**). When we performed Sanger sequencing on the product from these reactions, we observed a 100% concordance between the published SARS-CoV-2 viral sequence (GenBank: MT072688.1) and our PCR products (**Figure 4B**). Next, we examined our test’s sensitivity using SYBR RT-PCR (**Figure 4C**). Results showed that our assay’s limit for reliable detection is 10 viral copies per reaction. We used a Cq value of 35 cycles as the cut off for reliable detection. We observed that we could not reliably and reproducibly detect 1 or fewer viral copies per reaction. This suggests that our test would work best on samples with higher viral loads and might, in theory, miss cases where the disease is at its initial stage and the viral load is still relatively low.

**Figure 4.**
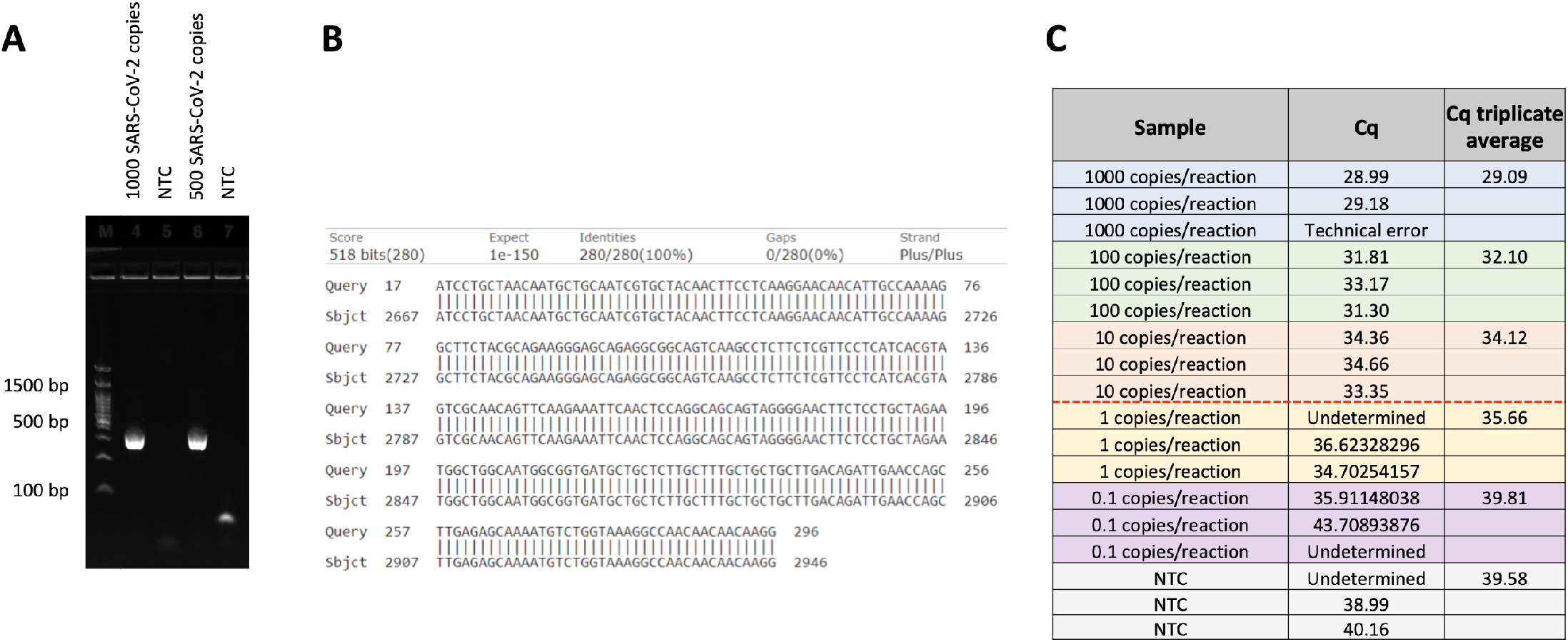
Specificity and sensitivity of a dual SARS-CoV-2 detection method relying on SYBR RT-PCR and Sanger sequencing. (A) Gel electrophoresis with PCR results from titrated positive and negative control samples using a primer pair amplifying a 328 bp region of the SARS-CoV-2 genome. (B) Nucleotide alignment between the published N gene SARS-CoV-2 sequence (Query) and a Sanger sequenced PCR product from the experiment in (A) marked as (Sbjct). (C) SYBR RT-PCR testing the sensitivity of the assay to different SARS-CoV-2 viral loads. Feline cDNA was spiked-in with titrated amounts of SARS-CoV-2 positive control plasmid. The red dashed line indicates the limit of reliable and reproducible detection.

We next tested our assay with two in-house RNA positive control samples obtained from an anonymous source. Each of them had a known concentration of 100 viral copies per reaction. We transcribed cDNA from these samples and ran our conventional PCR assay in order to perform Sanger sequencing on the resultant products. We observed a 100% concordance between the SARS-CoV-2 positive samples’ viral sequences and the published SARS-CoV-2 viral genome sequence (**Figure 5**). While preliminary, these results suggest that our assay could be used in a real-world scenario to identify SARS-CoV-2 positive feline and, potentially, human patients.

**Figure 5.**
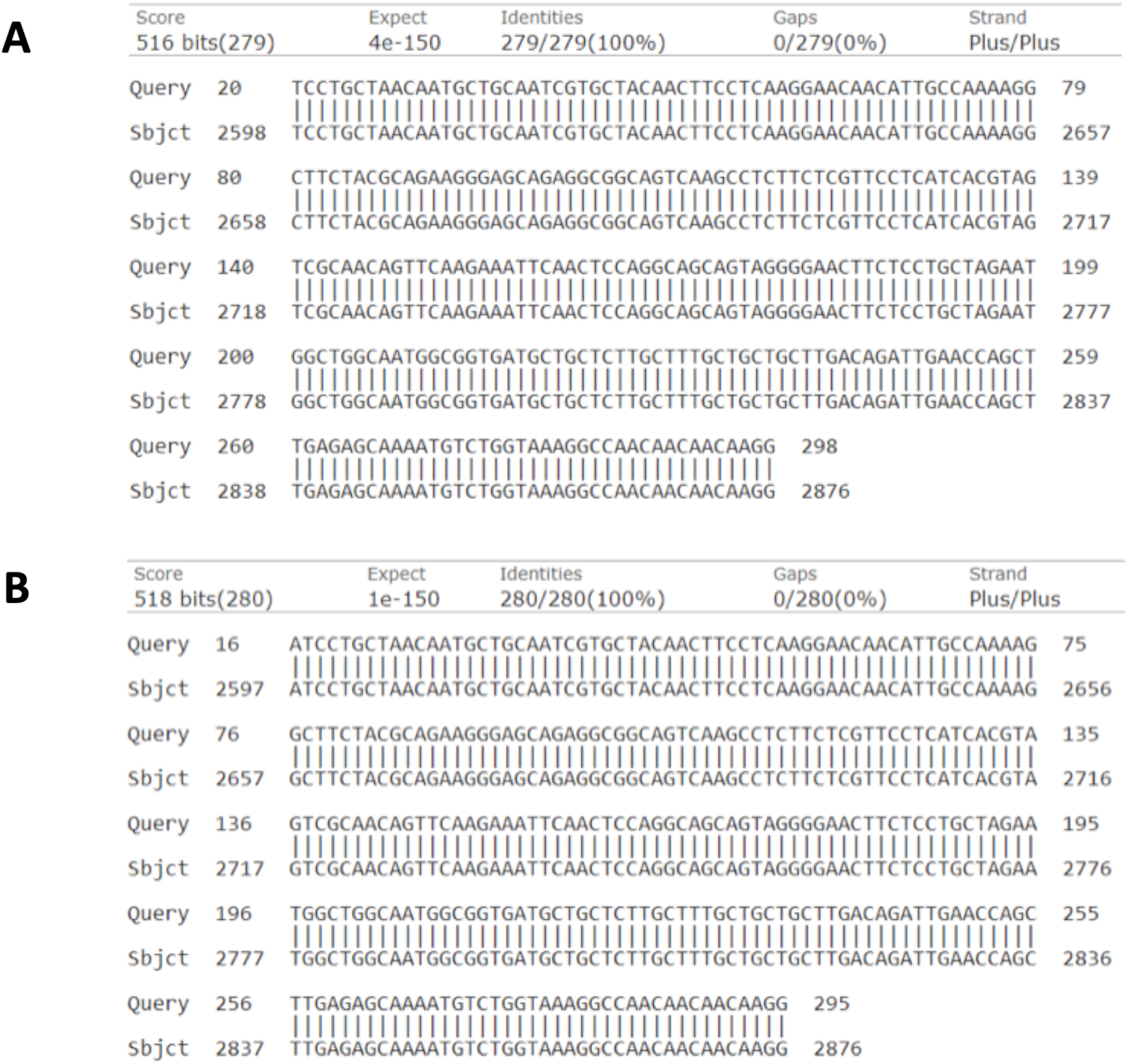
Nucleotide alignment between the published N gene SARS-CoV-2 sequence (Sbjct) and Sanger sequenced PCR products generated with our assay’s primers using two SARS-CoV-2 RNA positive control samples (A,B). The Sanger sequenced PCR products are marked as ‘Query’.

## Discussion

We developed a dual method PCR based test for the detection of SARS-CoV-2 in cats that does not rely on commonly used reagents and can, therefore, be used without significantly impacting the already strained COVID-19 testing reagent supply chain. Our assay is versatile and can be adapted to different laboratory setups and available reagents. While our test can be used with oropharyngeal collection kits designed specifically for RNA storage and stabilization, we demonstrated that some collection kits optimized for DNA preservation, such as the PERFORMAgene DNA PG-100 by DNAGenotek, have the potential to be used to collect and preserve feline and viral RNA. In addition, we demonstrated that an inexpensive, previously published^8^, SPRI beads-based RNA extraction method can be used for the successful extraction of total RNA from feline samples collected with ORAcollect RNA OR-100 (optimized for RNA stabilization) or PERFORMAgene DNA PG-100. One potential caveat is that, since long-term RNA preservation with the PERFORMAgene DNA PG-100 was not tested, samples collected in this manner should probably undergo rapid processing (within 24 hours) in order to maximize chances of successful viral detection.

During the COVID-19 pandemic, RT-PCR machines and TaqMan probe-based RT-PCR reagents can be costly and in short supply. As an alternative, we developed an assay that relies on SYBR RT-PCR and Sanger sequencing. Each of those two methodologies can be used in isolation, depending on preference and the purpose of testing. If the test is used in COVID-19 outbreak areas, the quicker SYBR RT-PCR version can be employed (results within 1 day), which does not distinguish between SARS-CoV and SARS-CoV-2. The rationale in such a scenario is that it is highly unlikely for SARS-CoV to be present in a COVID-19 affected area. For a more definitive answer where a clear distinction between SARS-CoV and SARS-CoV-2 is required, the Sanger sequencing version of our assay should be used alone or in combination with the SYBR RT-PCR (results within 2 days).

Our test’s sensitivity is 10 viral copies per reaction. Human patients’ SARS-CoV-2 viral loads in nasopharyngeal and oropharyngeal samples typically vary between 1.9 and 8 log10 RNA copies/ml^9^, which corresponds to a range between 0.395 and >5000 viral copies per reaction volume as defined in our assay. While not much data is available on the comparative viral loads in COVID-19 positive animal patients, these results suggest that our test is likely more suitable for animals that are symptomatic and are thus expected to have a higher viral load.

Although more studies are needed, our test could, in theory, be employed in human COVID-19 screening. The incorporation of Sanger sequencing in our assay has one significant advantage over RT-PCR diagnostics - single nucleotide resolution. This property could be exploited to catalog SARS-CoV-2 positive samples and look for strain-specific mutations in the N gene. This will allow a more accurate strain-level classification of positive cases without resorting to the much slower and more expensive whole viral genome sequencing. Using our method, different mutation hotspot regions of the SARS-CoV-2 genome can be targeted for amplification to better understand strain diversity and penetration in different populations.

## Materials and methods

### Oropharyngeal swab collection

We tested two buccal swab collection kits - ORAcollect RNA OR-100 and PERFORMAgene DNA PG-100, both available from DNAGenotek. While neither kit is designed specifically for oropharyngeal sample collection, we found it relatively straightforward to use either of them to collect an oropharyngeal swab sample from a cat. Samples were collected in the same sitting, from the same cat, with both collection kits. The cat had not had anything to eat or drink for an hour prior to sample collection.

### RNA extraction

We used a previously described method for purifying nucleic acids by solid-phase reversible immobilization (SPRI)^8,10^, with some protocol variations. The following is a brief protocol outline. First, each sample was heated at 55°C for 1 hour. The sample collected with the ORAcollect RNA OR-100 kit was then heated for 10 min at 90°C and the pH was adjusted to neutral, as per manufacturer’s instructions. This step was omitted for the sample collected with the PERFORMAgene DNA PG-100. Next, 2X volume of SPRI beads mix (Magnetic Beads Carboxylate MBC-200 from MCLAB) was added to each sample, mixed and incubated for 5 min prior to bead immobilization with a magnet. The bead pellet was washed twice with 80% freshly prepared ethanol and the RNA was eluted in 1XTE. Each sample was then treated with DNAse. EDTA, 3M Sodium acetate (pH 5.5) and 100% ethanol were used for RNA precipitation for 30 min at -20°C. Samples were then pelleted, washed with 70% ethanol and resuspended in 1XTE.

### cDNA synthesis

We used the iScript Select cDNA Synthesis Kit (1708896) from BioRad and followed manufacturer’s instructions.

### Custom primers

We designed the following primer pair amplifying 328 bp of the N region of the SARS-CoV-2 genome:

COVID_FP: CAACTGAGGGAGCCTTGAATAC

COVID_RP: CCTTGTTGTTGTTGGCCTTTAC

### RT-PCR

We used AppliedBiosystems’ PowerUp™ SYBR™ Green Master Mix (A25742) and followed manufacturer’s instructions for setting up 10 μl reactions. All consumables and pipettes were placed in a UV hood for 10 minutes prior to reaction setup. The RT-PCR program used was as follows: 1 × 2 min at 50°C, 1 × 2 min at 95°C, 50 X (15 sec at 95°C, 15 sec at 60°C, 1 min at 72°C). Reactions were performed in triplicate. Cat kidney total RNA (Zyagen) was used as a positive control in the RT-PCR experiments assessing the presence of feline housekeeping genes in the extracted RNA. The 2019-nCoV_N_Positive control #10006625 (IDT) was used as a spike-in control in RT-PCR reactions.

### RNA-seq

The KAPA RNA HyperPrep Kit with RiboErase (08098131702) was used for preparation of RNA-seq libraries. Manufacturer’s instructions were followed.

### Viral genome detection in RNA-seq dataset

To detect viruses from RNA-seq data, we used VirDetect^11^ (https://github.com/dmarron/virdetect). VirDetect begins by aligning RNA-seq reads to the feline genome^12^ using the STARv2.4 aligner^13^. Reads that did not align to the feline genome were then mapped to a viral genomes database optimized to increase specificity by masking the viral genomes for areas of feline homology and areas of low complexity.

## Data Availability

The authors confirm that the data supporting the findings of this study are available within the article.

## Notes

### Competing Interest Statement

The authors have declared no competing interest.

### Funding Statement

The work was supported by internal Basepaws funding. No external funding was utilized.

